# A comparison of the association of fat-to-muscle ratio vs. BMI and hypertension: A US population-based study

**DOI:** 10.64898/2025.12.14.25342233

**Authors:** Olubunmi O Oladunjoye, Dulcie Kermah, Keith Norris, Bettina M. Beech

## Abstract

**Objective:** Hypertension is the leading cause of preventable cardiovascular disease (CVD) and is associated with obesity. Body mass index (BMI), the most common measure of obesity, does not distinguish between fat and muscle mass. By contrast, dual-energy X-ray absorptiometry (DXA) is the gold standard for measuring fat composition with high accuracy and minimal variability. To our knowledge, no United States studies have examined the association between BMI or fat-to-muscle-ratio (FTMR) (using DXA) and hypertension.

**Methods:** We analyzed NHANES data of adults aged 20-59 years from 5 consecutive cycles (2011-2012 through 2017-2018). The primary outcome was hypertension. Logistic regression was used to determine the associations between FTMR and odds of hypertension, and between BMI and hypertension.

**Results:** The mean FTMR was higher in the hypertension compared to the non-hypertension group (0.57 ± 0.01 vs. 0.53 ± 0.004, p < 0.001), with a similar pattern observed for BMI (32.3 ± 0.2 vs. 28.1 ± 0.1, p < 0.001). Logistic regression analysis showed each one-unit increase in FTMR was associated with 2.80 times higher odds of hypertension, while each 1 kg/m² increase in BMI was associated with 1.08 times higher odds. The sex specific odds ratio (OR) for FTMR were even higher after adjusting for age (males 24.58 (11.74-51.46), females 8.77 (5.04-15.26), p<0.001). However, after adjusting for sex in a receiver operator curve (ROC) analysis, FTMR (Area Under the Curve [AUC] 0.63; 95% CI 0.62–0.64) did not outperform BMI (AUC 0.67; 95% CI 0.66–0.68) regarding their association with hypertension.

**Conclusion:** Although logistic regression showed a stronger relation of FTMR than BMI and the odds of hypertension, the ROC curve indicated no difference in association of hypertension and FTMR and BMI. Further research should examine the ability of FTMR compared to BMI to predict hypertension and CVD related-complications in high-risk subgroups.

## Introduction

Obesity is associated cardiometabolic diseases, including but not limited to hypertension, the leading cause of preventable cardiovascular disease[1–3]. Hypertension increases the risk for heart disease and stroke, which are two of the leading causes death in the United States (U.S.) [4]. Over 40% of U.S. adults 20 years and older have obesity and about 58% of U.S. adults with obesity have hypertension[5]. Treating obesity can lead to significant reductions in blood pressure; therefore, it is important to identify populations with increased adiposity for appropriate counseling, treatment, and engagement in self-management practices[6, 7].

Body mass index (BMI) remains the most widely utilized metric for assessing obesity in clinical practice; however, it lacks the ability to differentiate between adipose tissue and lean muscle mass. This leads to potential over- or underestimation of adiposity[8–12]. An underestimation of adiposity may lead to a lack of optimal counseling and treatment, while overestimation can lead to misattribution of symptoms, potentially delaying or misdirecting treatment for conditions unrelated to excess weight. More precise measures of obesity beyond BMI are necessary to accurately identify individuals with elevated adiposity who are at increased risk for obesity-related complications, including hypertension. Improved assessment tools can facilitate earlier detection and targeted intervention, ultimately helping to reduce the incidence of hypertension and its associated cardiovascular morbidity and mortality.

Alternative measures of adiposity, such as waist circumference, have been extensively studied and demonstrate strong correlations with metabolic diseases. However, waist circumference does not consistently reflect visceral fat and may be subject to inter-individual variability in measurement technique. Its predictive value for additional metabolic risk is also limited in individuals with a BMI greater than 35 kg/m², where excess adiposity may obscure meaningful distinctions in fat distribution [13]. Recent studies have focused on fat-to-muscle ratio (FTMR) to explore its association with obesity-related conditions, using methods such as skinfold calipers, air displacement plethysmography, and bioelectrical impedance analysis (BIA) [14–16]. However, these techniques are generally less accurate compared to computerized tomography (CT), magnetic resonance imaging (MRI), and dual-energy X-ray absorptiometry (DXA) [17].

Dual-energy X-ray absorptiometry (DXA) is considered the gold standard for accurately assessing body fat composition, offering high precision with minimal variability. A study by Seo et al. demonstrated that a high FTMR, as measured by DXA, is significantly associated with metabolic syndrome and insulin resistance in Korean adults [16]. However, there is currently no established consensus on the optimal FTMR cutoff value to define excess adiposity, as it may vary by age and sex. To date, no studies have specifically investigated the relationship between FTMR measured by DXA and hypertension within a U.S. population. Therefore, the objective of this study was to evaluate the association of hypertension prevalence with BMI and with FTMR as assessed by DXA.

## Materials and Methods

A secondary analysis of National Health and Nutrition Examination Survey (NHANES) data was conducted on adults aged 20-59 years from 5 consecutive cycles (2011-2012 through 2017-2018). The NHANES database is a cross-sectional survey conducted by the National Center for Health Statistics of the Centers for Disease Control and Prevention to assess the health and nutritional status of adults and children in the United States[18].

The primary outcome was hypertension, defined as a self-reported history of diagnosed hypertension, current use of antihypertensive medication or those with mean blood pressure reading greater than 140/90 mmHg. The primary independent variable was the fat-to-muscle ratio, calculated using fat and lean mass measurements obtained from whole-body DXA scans in the NHANES examination data. NHANES examinations include whole-body DXA scans performed only on participants aged 8 to 59 years, excluding all pregnant females. Individuals who self-reported radiographic contrast use within 7 days of the exam, or who weighed over 300 lbs. or were taller than 6’5”, were also excluded[18]. Demographic data (age, sex, race and ethnicity), socioeconomic factors (education, income), and other measurements such as BMI, were obtained from the NHANES datasets. Participant race and ethnicity were self-reported. A complete case analysis was done, i.e. all participants with missing information on age, gender/ethnicity and hypertension were excluded from the analysis (figure 1). The fat-to-muscle ratio was divided into quartiles and the prevalence of hypertension within each quartile was determined. This was also performed within each BMI category of normal weight, overweight, and obesity.

**Fig 1:**
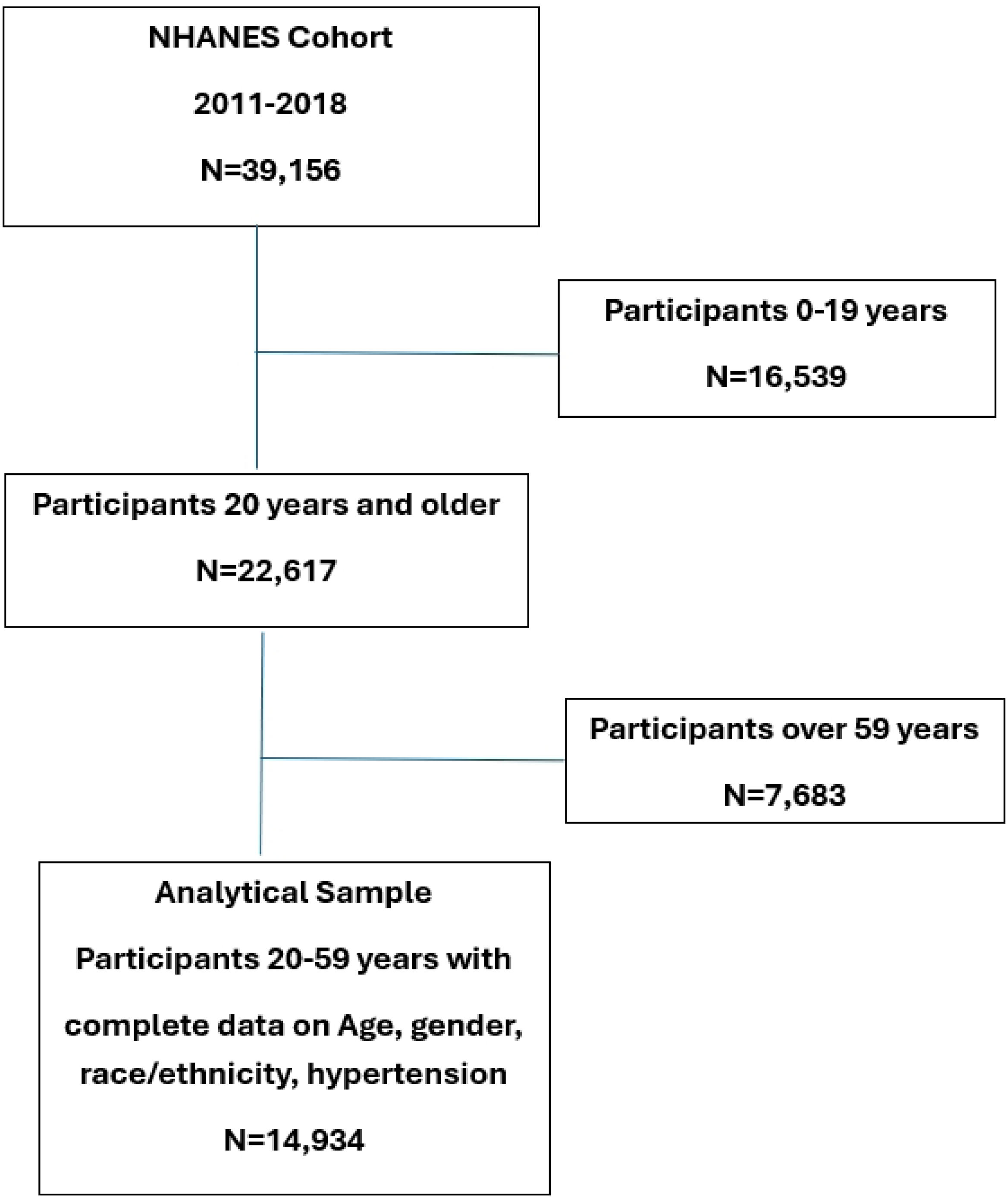
Flow Diagram of Participant.

Continuous variables were reported as means ± standard deviations, or as medians with interquartile ranges for skewed data. Categorical variables were presented as proportions and/or percentages. Logistic regression was used to determine the association between FTMR and odds of hypertension as well as the association between BMI and odds of prevelant hypertension. BMI and FTMR and their association with hypertension were compared using the receiver operating characteristic (ROC) analysis and area under the curve calculation. P<0.05 (two-sided) was considered statistically significant. Statistical analyses were performed using SAS version 9.4 (SAS Institute, Cary, NC, USA).

## Results

### Characteristics of Study population

The mean age of participants was 39.5 ± 0.2 years. Individuals with hypertension were significantly older than those without the condition, with mean ages of 45 ± 0.2 and 37.1 ± 0.2 years, respectively (p < 0.001). Hypertension was more prevalent among males (30.9%) compared to females (24.8%) (p < 0.001). Additionally, the hypertensive group exhibited higher mean BMI values (32.3 ± 0.2 vs. 28.1 ± 0.1; p < 0.001) and FTMR levels (0.57 ± 0.01 vs. 0.53 ± 0.004; p < 0.001).

The average age of male and female participants was comparable. The mean BMI was slightly lower in males (29.0 kg/m²) than in females (29.5 kg/m²), with the difference reaching statistical significance (p < 0.001). Mean FTMR values differed markedly between sexes, with males averaging 0.40 ± 0.01 and females 0.68 ± 0.01. Comprehensive demographic characteristics stratified by hypertension status and sex are presented in Tables 1 and 2, respectively.

**Table 1:** Demographic characteristics of participants with and without hypertension.

| Variables | Total N (%) | Hypertension | No hypertension | p-value |
| --- | --- | --- | --- | --- |
| Age (mean±SE) | 39.5 ($\pm 0.2$) | 45.0($\pm 0.2$) | 37.1 ($\pm 0.2$) | <0.001 |
| • 20-39 years | 7211 (49.9) | 1133 (7.7) | 6078 (42.2) |  |
| • 40-59 years | 7136 (50.1) | 3042 (20.0) | 4094 (30.0) |  |
| Sex |  |  |  | <0.001 |
| • Male | 6848 (49.1) | 2141 (30.9) | 4707 (69.1) |  |
| • Female | 7499 (50.9) | 2034 (24.8) | 5465 (75.2) |  |
| Race |  |  |  | <0.001 |
| • White | 4826 (60.3) | 1388 (27.9) | 3438 (72.1) |  |
| • Black | 3271 (12.3) | 1296 (37.9) | 1975 (62.1) |  |
| • Hispanic | 3528 (17.6) | 865 (22.5) | 2673 (77.5) |  |
| • Other | 2712 (9.7) | 626 (23.8) | 2086 (76.2) |  |
| Education |  |  |  | 0.003 |
| • Less than High School | 2762 (13.7) | 885 (30.4) | 1877 (69.6) |  |
| • High School | 3116 (21.9) | 1006 (31.2) | 2110 (68.8) |  |
| • Beyond High School | 8465 (64.5) | 2284 (26.0) | 6181 (74.0) |  |
| Marital Status |  |  |  | <0.001 |
| • Married/living with partner | 8580 (62.4) | 2482 (28.3) | 6098 (71.7) |  |
| • Divorced/separated | 1839 (12.1) | 790 (40.9) | 1049 (59.1) |  |
| • Widowed | 200 (1.1) | 92 (43.8) | 108 (56.2) |  |
| • Never married | 3725 (24.4) | 811 (19.1) | 2914 (80.9) |  |
| BMI |  |  |  | <0.001 |
| Mean BMI (mean±SE) |  | 32.3 (±0.2) | 28.1 (±0.1) |  |
| • Normal | 4090 (29.0) | 649 (14.2) | 3441 (85.8) |  |
| • Overweight | 4316 (31.7) | 1125 (25.4) | 3191 (74.6) |  |
| • Obesity | 5549 (39.3) | 2307 (40.0) | 3242 (60.0) |  |
| FTMR (mean±SE) | 0.54 ± 0.004 | 0.57 (±0.01) | 0.53 (±0.004) |  |
| • FTMR <0.38 | 2756 (25.2) | 562 (4.8) | 2184 (20.4) | <0.001 |
| • FTMR 0.38-0.49 | 2533 (24.2) | 749 (7.3) | 1784 (16.9) |  |
| • FTMR 0.50-0.68 | 2804 (25.9) | 787 (6.7) | 2017 (19.2) |  |
| • FTMR ≥0.69 | 2781 (24.7) | 959 (7.8) | 1822 (16.9) |  |

BMI: Body mass index; FTMR: Fat to muscle ratio

**Table 2:** Demographic characteristics of participants with and without hypertension by Sex.

|  | Males |  |  |  | Females |  |  |  |
| --- | --- | --- | --- | --- | --- | --- | --- | --- |
| Variables | Total N (%) | Hypertension | No hypertension | p-value | Total N (%) | Hypertension | No hypertension | p-value |
| Age (mean±SE) | 39.4 (±0.2) | 45.0 (±0.3) | 36.9 (±0.3) | <0.001 | 39.6 (± 0.3) | 46.5 (±0.3) | 37.3 (±0.3) | <0.001 |
| • 20-39 years | 3491 (50.3) | 654 (18.9) | 2837 (81.1) |  | 3729 (49.5) | 479 (12.1) | 3241 (87.9) |  |
| • 40-59 years | 3357 (49.7) | 1487 (43.0) | 1870 (57.0) |  | 3779 (50.5) | 1555 (37.2) | 2224 (62.8) |  |
| Race |  |  |  | <0.001 |  |  |  | <0.001 |
| • White | 2376 (60.7) | 774 (32.1) | 1602 (67.9) |  | 2450 (59.9) | 614 (23.7) | 1836 (76.3) |  |
| • Black | 1503 (11.4) | 600 (38.2) | 903 (62.0) |  | 1768 (13.2) | 696 (37.6) | 1072 (62.4) |  |
| • Hispanic | 1631 (18.1) | 415 (24.3)) | 1216 (75.7) |  | 1907 (17.1) | 450 (20.6) | 1457 (79.4) |  |
| • Other | 1388 (9.7) | 352 (26.7) | 986 (73.3)) |  | 1374 (9.8) | 274 (21.1) | 1100 (78.9) |  |
| Education |  |  |  | 0.058 |  |  |  | <0.001 |
| • Less than High School | 1444 (15.2) | 452 (28.6) | 992 (71.4) |  | 1318 (12.2) | 433 (32.6) | 885 (67.4) |  |
| • High School | 1634 (24.3) | 555 (33.5) | 1079 (66.5) |  | 1482 (19.5) | 451 (28.4) | 1031 (71.6) |  |
| • Beyond High School | 3770 (60.5) | 1134 (30.4) | 2636 (69.6) |  | 4695 (68.2) | 1150 (22.3) | 3545 (77.7) |  |
| Marital Status |  |  |  | <0.001 |  |  |  | <0.001 |
| • Married/living with partner | 4199 (62.4) | 1371 (32.5) | 2828 (67.5) |  | 4381 (62.4) | 1111 (24.2) | 3270 (75.8) |  |
| • Divorced/separated | 688 (10.0) | 330 (47.5) | 358 (52.5) |  | 1151 (14.2) | 460 (36.4) | 691 (63.6) |  |
| • Widowed | 49 (0.6) | 21 (39.3) | 28 (60.7) |  | 151 (1.6) | 71 (45.5) | 80 (54.5) |  |
| • Never married | 1910 (27.0) | 419 (20.7) | 1491 (79.3) |  | 1815 (21.8) | 392 (17.4) | 1423 (82.6) |  |
| Mean BMI (mean±SE) | 29.0 (±0.2) | 31.4 (±0.2) | 28.0 (±0.2) | <0.001 | 29.5 (±0.2) | 33.4 (±0.3) | 28.3 (±0.2) | <0.001 |
| • Normal | 1879 (25.9) | 337 (16.4) | 1542 (83.6) |  | 2211 (32.0) | 312 (12.5) | 1899 (87.5) |  |
| • Overweight | 2393 (36.2) | 695 (28.9) | 1698 (71.1) |  | 1923 (27.3) | 430 (21.0) | 1493 (79.0) |  |
| • Obesity | 2412 (37.9) | 1065 (43.1) | 1347 (56.9) |  | 3137 (40.7) | 1242 (37.1) | 1895 (62.9) |  |
| FTMR (mean±SE) | 0.40 (±0.01) | 0.44 (±0.00) | 0.38 (±0.00) |  | 0.68 (±0.01) | 0.74 (±0.01) | 0.66 (±0.01) | <0.001 |
| • FTMR <0.38 | 2555 (46.2) | 541 (19.9) | 2014 (80.1) | <0.001 | 191 (4.0) | 21 (10.2) | 170 (89.8) |  |
| • FTMR 0.38-0.49 | 1900 (35.3) | 663 (36.1) | 1237 (63.9) |  | 633 (13.0) | 86 (14.4) | 547 (85.6) |  |
| • FTMR 0.50-0.68 | 843 (16.2) | 358 (41.5) | 485 (58.5) |  | 1961 (35.3) | 429 (18.3) | 1532 (81.7) |  |
| • FTMR ≥0.69 | 104 (1.9) | 46 (40.6) | 58 (59.4) |  | 2677 (47.7) | 913 (31.3) | 1764 (68.7) |  |

BMI: Body Mass Index; FTMR: Fat to muscle ratio

Table 2 presents demographic characteristics of participants, with and without hypertension stratified by sex. Among males, those with hypertension had a significantly higher mean age of 45 (±0.3) years compared to 36.9 (±0.3) years for those without hypertension (p<0.001). Hypertension prevalence varied by race, with 38.2% of Black males affected, followed by 32.1% of White males, 26.7% of males from other races (p<0.001), and 24.3% of Hispanic males. Hypertensive males also had a higher mean BMI (31.4 ±0.2 kg/m2) than non-hypertensive males (28.0 ±0.2kg/m2) (p<0.001). The prevalence of hypertension increased across BMI categories: 16.4% among males with normal BMI, 28.9% among those overweight, and 43.1% among those with obesity. Similarly, hypertension prevalence rose with increasing with FTMR levels, from 19.9% in males with FTMR <0.38 to 40.6% for FTMR ≥0.69

As observed in males, females with hypertension were significantly older than those without the condition, with mean age 46.5 ±0.3 years and 37.3 ±0.3 years, respectively (p-value <0.001). Among racial groups, Black females exhibited the highest prevalence of hypertension at 37.6%. Hypertensive females also had significantly higher mean (33.4 ±0.3 kg/m2) compared to non-hypertensive (28.3 ±0.2kg/m2) (p<0.001). The prevalence of hypertension increased across BMI categories: 12.5% among females with normal BMI, 21.0% among those overweight, and 37.1% among those with obesity. A similar trend was observed with FTMR levels, with hypertension prevalence rising from10.2% in females with FTMR <0.38 to 31.3% for FTMR ≥0.69

### Association between the presence of hypertension and both FTMR and BMI

Table 3 summarizes the associations between hypertension and both FTMR and BMI in male and female participants. Across the total population – and within each sex subgroup - FTMR and BMI emerged as statistically significant predictors of hypertension. Notably, the odds ratios (ORs) for FTMR were consistently higher than those for BMI, underscoring its stronger predictive value.

**Table 3:** Association between FTMR vs BMI and presence of hypertension.

| Variable |  | Unadjusted OR | p-value | Adjusted OR (age) | p-value |
| --- | --- | --- | --- | --- | --- |
| FTMR | Total | 2.80 (2.13-3.70) | <0.001 | 1.79 (1.34-2.90) | <0.001 |
|  | Male | 37.14 (19.0-73.0) | <0.001 | 24.58 (11.74-51.46) | <0.001 |
|  | Female | 14.76 (8.78-24.82) | <0.001 | 8.77 (5.04-15.26) | <0.001 |
| BMI | Total population | 1.08 (1.07-1.09) | <0.001 | 1.08 (1.07-1.09) | <0.001 |
|  | Male | 1.09 (1.08-1.10) | <0.001 | 1.09 (1.07-1.10) | <0.001 |
|  | Female | 1.08 (1.07-1.09) | <0.001 | 1.08 (1.07-1.09) | <0.001 |

BMI: Body mass index; FTMR: Fat to muscle ratio; OR: Odds ratio Adjusted for age

FTMR was categorized into quartiles: (Q1) (< 0.38), Q2 (0.38–0.49), Q3 (0.50–0.68), and Q4 (≥ 0.69), with Q1 serving as the reference group. Among males, adjusted odds of hypertension increased progressively across quartiles, indicating a clear dose-response relationship. The other quartiles were compared with the first quartile. Compared to Q1, males in Q2 had 1.91 times the adjusted odds of hypertension (95% CI: 1.57-2.33, p<0.001 while Q3 had 2.54 times the adjusted odds (95% CI: 2.00-3.21, p<001). Males in Q4 had the highest adjusted odds, at 2.73 times (95% CI: 1.36-5.48, p=0.001) (Table 4). This shows a strong “dose-response” relationship between higher FTMR and hypertension risk in males. In contrast, among females, only those in the highest FTMR quartile (Q4) showed a statistically significant increase in hypertension risk compared to Q1, with an OR of 2.45 (95% CI: 1.31-4.57, p=0.005).

**Table 4:** Association between FTMR in quartiles and Hypertension.

| Hypertension | FTMR Quartiles | Unadjusted OR (95% CI) |  | Adjusted OR (for age) | p-value | Adjusted for race and age |  |
| --- | --- | --- | --- | --- | --- | --- | --- |
| Male | Q2 vs Q1 | 2.27 (1.88-2.74) | <0.001 | 1.91 (1.57-2.33) | <0.001 | 2.00 (1.64-2.44) | <0.001 |
|  | Q3 vs Q1 | 2.84 (2.30 - 3.51) | <0.001 | 2.54 (2.00-3.21) | <0.001 | 2.62 (2.06 – 3.34) | <0.001 |
|  | Q4 vs Q1 | 2.74 (1.60-4.70) | <0.001 | 2.73 (1.36-5.48) | 0.006 | 2.88 (1.42-5.83) | 0.004 |
| Female | Q2 vs Q1 | 1.47 (0.76-2.85) | 0.245 | 1.22 (0.61-2.44) | 0.562 | 1.25 (0.62 – 2.50) | 0.524 |
|  | Q3 vs Q1 | 1.96 (1.03-3.74) | 0.041 | 1.30 (0.67-2.53) | 0.434 | 1.28 (0.65 – 2.50) | 0.473 |
|  | Q4 vs Q1 | 3.99 (2.17-7.35) | <0.001 | 2.45 (1.31-4.57) | 0.006 | 2.39 (1.27 – 4.47) | 0.007 |
| Total population | Q2 vs Q1 | 1.83 (1.54-2.19) | <0.001 | 1.61 (1.34-1.93) | <0.001 | 1.68 (1.40-2.02) | <0.001 |
|  | Q3 vs Q1 | 1.46 (1.24 – 1.73) | <0.001 | 1.24 (1.04-1.47) | 0.019 | 1.28 (1.07-1.52) | 0.009 |
|  | Q4 vs Q1 | 1.95 (1.67-2.28) | <0.001 | 1.52 (1.29-1.79) | <0.001 | 1.54 (1.31-1.81) | <0.001 |

FTMR: Fat to muscle ratio; OR: Odds ratio; Q: Quartiles; Q1 <0.38, Q2 0.38- 0.49, Q3 0.50-0.68, Q4 ≥0.69;
Adjusted for age

In the BMI category, both overweight and obesity classifications are strongly and significantly associated with increased odds of hypertension across males, females, and the overall population, even after adjusting for age (Table 5).

**Table 5:** Association between BMI classification and Hypertension.

| Hypertension | BMI Category | Unadjusted OR (95% CI) | p-value | Adjusted OR for age | p-value | Adjusted for age and race |  |
| --- | --- | --- | --- | --- | --- | --- | --- |
| Male | Normal | reference |  |  |  |  |  |
|  | overweight | 2.07 (1.68-2.55) | <0.001 | 1.71 (1.38-2.12) | <0.001 | 1.78 (1.42-2.21) | <0.001 |
|  | obesity | 3.86 (3.08-4.86) | <0.001 | 3.41 (2.72-4.28) | <0.001 | 3.53 (2.81-4.44) | <0.001 |
| Female | Normal | reference |  | - |  |  |  |
|  | overweight | 1.85 (1.50-2.83) | <0.001 | 1.68 (1.35-2.09) | <0.001 | 1.66 (1.33-2.07) | <0.001 |
|  | obesity | 4.12 (3.43-4.95) | <0.001 | 3.99 (3.31-4.80) | <0.001 | 3.81 (3.15-4.59) | <0.001 |
| Total population | Normal | Reference |  |  |  |  |  |
|  | overweight | 2.05 (1.77-2.38) | <0.001 | 1.79 (1.54 – 2.07) | <0.001 | 1.82 (1.56-2.12) | <0.001 |
|  | obesity | 4.01 (3.41-4.72) | <0.001 | 3.72 (3.18-4.36) | <0.001 | 3.71 (3.16-4.37) | <0.001 |

BMI: Body mass index; OR: Odds ratio; Adjusted for age

### ROC Curve analysis

After adjusting for sex in an ROC analysis, FTMR (AUC 0.63; 95% CI 0.62–0.64) did not outperform BMI (AUC 0.67; 95% CI 0.66–0.68) in its association with hypertension (figures 2 and 3)

**Fig 2:**
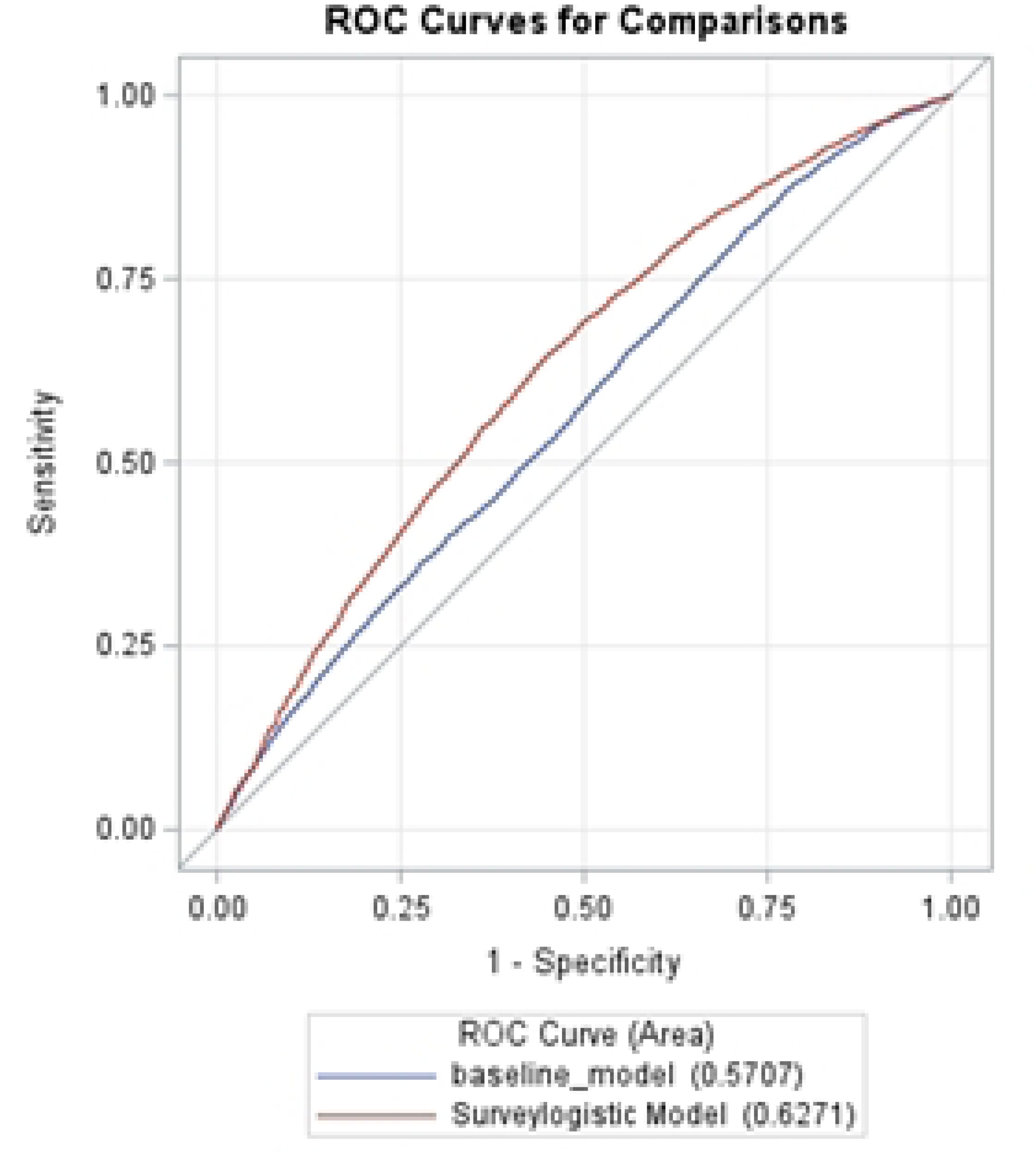
ROC Curve for association of FTMR and hypertension.

**Fig 3:**
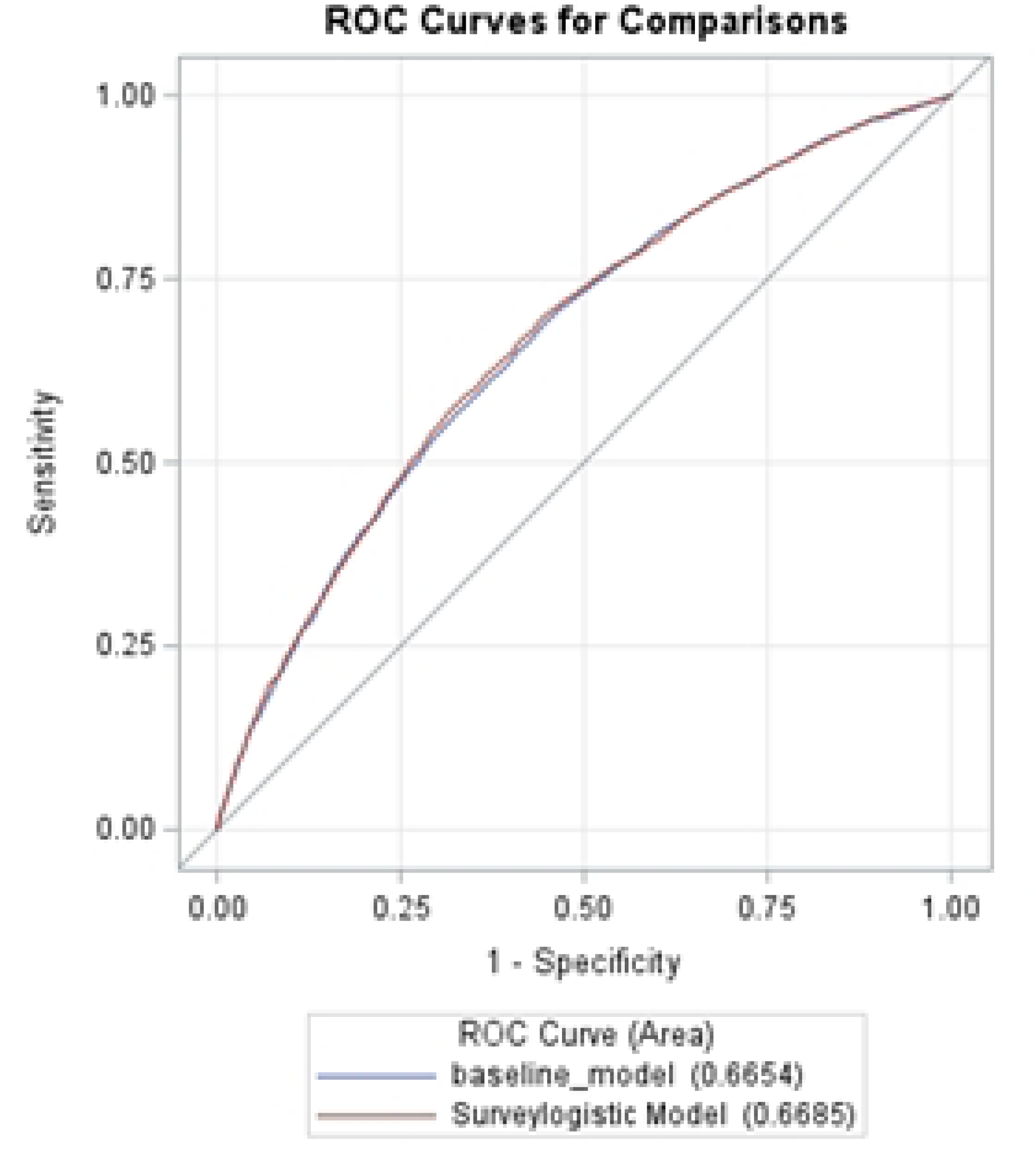
ROC Curve for association of BMI and hypertension.

## Discussion

This cross-sectional study utilized NHANES data from adults aged 20-59 years from 5 consecutive cycles (2011-2012 through 2017-2018) to compare the association of hypertension with FTMR and BMI. This study demonstrated that individuals with hypertension had significantly higher mean FTMR and BMI values compared to those without hypertension, a pattern that persisted across both sexes. Furthermore, when analyzed as continuous variables, FTMR showed a stronger association with hypertension than BMI. After adjusting for age, the adjusted ORs for FTMR were substantially higher at 24.58 for males and 8.77 for females, compared to BMI which yielded ORs of 1.09 for males, 1.08 and for females.

The association between BMI and hypertension have been well documented in the literature. FTMR is an emerging measure that is considered in defining adiposity. Studies have shown the correlation between fat mass, muscle mass and FTMR with diabetes, metabolic syndrome, cardiometabolic diseases, and hypertension [15, 16, 19]. Our study not only looked at the association of FTMR or BMI and hypertension, but it also compared the association of FTMR and hypertension with that of BMI and hypertension. The higher OR for association for FTMR compared to BMI in its association with hypertension suggests that a unit increase in FTMR may be more predictive of hypertension risk than a unit increase in BMI when considered as continuous measures.

Both FTMR and BMI demonstrated a statistically significant higher hypertension risk across ascending categories. For BMI, the adjusted ORs rose significantly from the “Normal” to “Overweight” and “Obesity” classifications with significance maintained even when stratified by sex. Similarly, FTMR showed a progressive increase in hypertension risk across quartiles among males, indicating a strong relationship. In females, although the FTMR was a strong overall predictor, a statistically significant increase in hypertension risk was observed only in the highest quartile (Q4). These findings aligned with those by Yuan-Yuei Chen et al who reported that higher FTMR quartiles were more predictive of hypertension in both sexes. However, their study employed sex-specific FTMR cutoffs, which differed from the uniform quartile approach used in the present analysis [15]. A recent study conducted in China by Zhe Chen et al also demonstrated that the risk of hypertension increased across FTMR quartiles. Interestingly, their findings showed a more pronounced relation in females (OR 6.663, 95%CI 2.652–16.742) than in males (OR: 2.647, 95%CI 1.740–4.027), which contrasts with the results of our study where the association was stronger in males[20]. This differences may be due to national cultural differences in diet and exercise modifying the relationship between obesity and hypertension.

This study has several strengths, including the use of a large, representative national sample. However, it also has limitations. NHANES’s body composition assessment was restricted to those younger than age 59 years. Cross sectional design limits the ability to determine causal relationships between variables. Patient’s hypertension status include participants self-report. To reduce classification bias,.we also assessd hypertension by current use of antihypertensive medication or those with mean blood pressure reading greater than 140/90 mmHg

## Conclusion

Both FTMR and BMI are strongly associated with an increased likelihood of hypertension, and both measures of obesity are significantly higher in individuals with hypertension. While BMI provides a consistently clear and significant gradient of risk across its classification categories for both sexes, FTMR, particularly as a continuous variable, demonstrates an even greater magnitude of association, suggesting it may be a very powerful predictor, especially for males across the weight spectrum and at higher FTMR in females.

## Data Availability

The data used in the study are publicly available from the Centers for Disease Control and Prevention

https://wwwn.cdc.gov/nchs/nhanes/

## Notes

### Competing Interest Statement

The authors have declared no competing interest.

### Funding Statement

Yes

### Author Declarations

This project involves secondary analysis of publicly available, de-identified data from the National Health and Nutrition Examination Survey (NHANES). The NHANES datasets are released by the National Center for Health Statistics (NCHS) with all personal identifiers removed and are fully compliant with federal standards for public-use data. Because the dataset contains no identifiable private information and the research involves analysis of existing, publicly available data, this project meets the criteria for exempt human subjects research under 45 CFR 46.104(d)(4).

## References

1. Feyisa BR, Tamiru A, Debelo S, Feyisa I, Tola EK, Tolesa EJ, et al. Magnitude of hypertension and its association with obesity among employees of Wallaga University, Ethiopia: a cross-sectional study. BMJ Open. 2023;13(7):e070656. Epub 2023/07/13. doi: 10.1136/bmjopen-2022-070656. PubMed PMID: 37438078; PubMed Central PMCID: PMCPMC10347519.

2. Mikhail N, Golub MS, Tuck ML. Obesity and hypertension. Prog Cardiovasc Dis. 1999;42(1):39–58. Epub 1999/10/03. doi: 10.1016/s0033-0620(99)70008-3. PubMed PMID: 10505492.

3. Global Burden of Metabolic Risk Factors for Chronic Diseases Collaboration. Cardiovascular disease, chronic kidney disease, and diabetes mortality burden of cardiometabolic risk factors from 1980 to 2010: a comparative risk assessment. Lancet Diabetes Endocrinol. 2014;2(8):634–47. Epub 2014/05/21. doi: 10.1016/s2213-8587(14)70102-0. PubMed PMID: 24842598; PubMed Central PMCID: PMCPMC4572741.

4. Xu J, Murphy S, Kockanek K, Arias E. Mortality in the United States, 2021. Hyattsville, MD: National Center for Health Statistics2022.

5. Stierman B, Afful J, Carroll MD, Chen T-C, Davy O, Fink S, et al. National Health and Nutrition Examination Survey 2017-March 2020 prepandemic data files-development of files and prevalence estimates for selected health outcomes. National health statistics reports. 2021;(158):10.15620/cdc:106273.

6. Martin SS, Aday AW, Almarzooq ZI, Anderson CA, Arora P, Avery CL, et al. 2024 heart disease and stroke statistics: a report of US and global data from the American Heart Association. Circulation. 2024;149(8):e347–e913.

7. Leggio M, Lombardi M, Caldarone E, Severi P, D’Emidio S, Armeni M, et al. The relationship between obesity and hypertension: an updated comprehensive overview on vicious twins. Hypertension Research. 2017;40(12):947–63. doi: 10.1038/hr.2017.75.

8. Khanna D, Peltzer C, Kahar P, Parmar MS. Body Mass Index (BMI): A Screening Tool Analysis. Cureus. 2022;14(2):e22119. Epub 2022/03/22. doi: 10.7759/cureus.22119. PubMed PMID: 35308730; PubMed Central PMCID: PMCPMC8920809.

9. Dalton M, Cameron AJ, Zimmet PZ, Shaw JE, Jolley D, Dunstan DW, et al. Waist circumference, waist-hip ratio and body mass index and their correlation with cardiovascular disease risk factors in Australian adults. J Intern Med. 2003;254(6):555–63. Epub 2003/12/04. doi: 10.1111/j.1365-2796.2003.01229.x. PubMed PMID: 14641796.

10. Sweatt K, Garvey WT, Martins C. Strengths and Limitations of BMI in the Diagnosis of Obesity: What is the Path Forward? Curr Obes Rep. 2024. Epub 2024/07/03. doi: 10.1007/s13679-024-00580-1. PubMed PMID: 38958869.

11. Rothman KJ. BMI-related errors in the measurement of obesity. Int J Obes (Lond). 2008;32 Suppl 3:S56–9. Epub 2008/08/21. doi: 10.1038/ijo.2008.87. PubMed PMID: 18695655.

12. Coutinho T, Goel K, Corrêa de Sá D, Kragelund C, Kanaya AM, Zeller M, et al. Central obesity and survival in subjects with coronary artery disease: a systematic review of the literature and collaborative analysis with individual subject data. J Am Coll Cardiol. 2011;57(19):1877–86. Epub 2011/05/07. doi: 10.1016/j.jacc.2010.11.058. PubMed PMID: 21545944.

13. Seagle HM, Wyatt HR, Hill JO. Chapter 24 - Obesity: Overview of Treatments and Interventions. In: Coulston AM, Boushey CJ, Ferruzzi MG, editors. Nutrition in the Prevention and Treatment of Disease (Third Edition): Academic Press; 2013. p. 445–64.

14. Eun Y, Lee SN, Song SW, Kim HN, Kim SH, Lee YA, et al. Fat-to-muscle Ratio: A New Indicator for Coronary Artery Disease in Healthy Adults. Int J Med Sci. 2021;18(16):3738–43. Epub 2021/11/19. doi: 10.7150/ijms.62871. PubMed PMID: 34790047; PubMed Central PMCID: PMCPMC8579285.

15. Chen YY, Fang WH, Wang CC, Kao TW, Yang HF, Wu CJ, et al. Fat-to-muscle ratio is a useful index for cardiometabolic risks: A population-based observational study. PLoS One. 2019;14(4):e0214994. Epub 2019/04/10. doi: 10.1371/journal.pone.0214994. PubMed PMID: 30964893; PubMed Central PMCID: PMCPMC6456204.

16. Xu K, Zhu HJ, Chen S, Chen L, Wang X, Zhang LY, et al. Fat-to-muscle Ratio: A New Anthropometric Indicator for Predicting Metabolic Syndrome in the Han and Bouyei Populations from Guizhou Province, China. Biomed Environ Sci. 2018;31(4):261–71. Epub 2018/05/19. doi: 10.3967/bes2018.034. PubMed PMID: 29773089.

17. Ceniccola GD, Castro MG, Piovacari SMF, Horie LM, Corrêa FG, Barrere APN, et al. Current technologies in body composition assessment: advantages and disadvantages. Nutrition. 2019;62:25–31. doi: 10.1016/j.nut.2018.11.028.

18. Centers for Disease Control and Prevention (CDC). National Center for Health Statistics (NCHS). National Health and Nutrition Examination Survey Data. Hyattsville, MD: U.S. Department of Health and Human Services, Centers for Disease Control and Prevention.

19. Brown CD, Higgins M, Donato KA, Rohde FC, Garrison R, Obarzanek E, et al. Body Mass Index and the Prevalence of Hypertension and Dyslipidemia. Obesity Research. 2000;8(9):605–19. doi: 10.1038/oby.2000.79.

20. Chen Z, Guo D, Xiao L, Su H, Chen Y. Association of fat-to-muscle ratio with hypertension: a cross-sectional study in China. Journal of Human Hypertension. 2025;39(4):301–7. doi: 10.1038/s41371-025-00992-z.

